# Effectiveness of the mRNA BNT162b2 vaccine six months after vaccination: findings from a large Israeli HMO

**DOI:** 10.1101/2021.09.01.21262957

**Authors:** Jennifer Kertes, Sharon Baruch Gez, Yaki Saciuk, Lia Supino-Rosin, Naama Shamir Stein, Miri Mizrahi-Reuveni, Anat Ekka Zohar

## Abstract

Israel is currently experiencing a new wave of CoVid-19 infection, six months after implementing a national vaccination campaign. We carried out three discrete analyses using data from a large Israeli HMO to determine whether IgG levels of those fully vaccinated drop over time, the relationship between IgG titer and subsequent PCR-confirmed infection, and compare PCR-confirmed infection rates by period of vaccination. We found that mean IgG antibody levels steadily decreased over the six-month period in the total tested population, and in all age groups. An inverse relationship was found between IgG titer and subsequent CoVid-19 infection (PCR-positive). Those participants vaccinated in the first two months of the campaign were more likely to become infected than those subsequently vaccinated. The 60+ vaccinated had lower initial IgG levels, and were at greater risk of infection. The findings support the decision to add a booster vaccine for those aged 60 and over.

**Article Summary Line:** The BNT162b2 vaccine was found to be less effective in protecting against Covid-19 infection after six months, and vaccination with a third dose is indicated.

## INTRODUCTION

The first case of Covid-19 was identified in Israel at the end of February 2019.*(1)* As in other countries, Israel has experienced a number of infection waves. The third and largest wave to-date, largely attributed to the entry of the Alpha variant to Israel, commenced in September 2020. At its’ peak, over 8,000 new cases were being identified daily.*(2)* Israel was among the first countries to introduce a national vaccination campaign, using the mRNA BNT162b2 vaccine (Pfizer-BioNTech©). The BNT162b2 vaccine received emergency approval for use by the FDA after demonstrating 95% efficacy over an average two-month follow-up period.*(3,4)* The vaccine was initially approved for any person aged 16 and over, with a recommended 21-day interval, two-dose administration. The vaccine campaign commenced on the 20^th^ December 2020 (concurrent with a two-month nationwide lockdown), first targeting all healthcare workers and the 60+ population, and quickly extending to all the population aged 16+. Initially those members with a prior infection were not eligible for vaccination, but within three months, policy was changed to offer a single dose to all those with prior infection. By April 2021, over 50% of those aged 16 and over, and 88% of the 50+ age group country-wide had been fully vaccinated.*(2)* Incidence rates dropped to 140/day by April 2021.*(2)* Initial population-based studies in Israel comparing the vaccinated and unvaccinated, reported vaccine effectiveness rates of 95%.*(5,6)*

One of the biggest questions regarding the vaccine is the length of protection provided. In publishing its third phase results, Pfizer-BioNTech reported a 91% efficacy rate over a six month follow-up period, with an estimated 6% drop in efficacy every two months.*(7)* Israeli population-based observational studies are no longer a feasible method of evaluating long-term effectiveness of the vaccine, given that the majority of the population have now been fully vaccinated. Infection rates in Israel are on the rise again, with Israel currently experiencing it’s fourth wave of infection. The majority (97%) of those infected in this current wave are infected with the Delta variant (B.1.617.2) (unpublished data provided by Israel Ministry of Health Laboratories). Initial serological studies of the Delta variant suggest that the BNT162b2 vaccine provides protection against Delta-variant infection, but at lower rates than for the Alpha variant (88% vs 93.7% respectively).*(8)* Given the rise in infection rates, the dilemma arose whether this rise was attributable to reduced effectiveness of the vaccine against the Delta variant or a waning of protection provided by the vaccine over time.

The objective of this study was to determine if the BNT162b2 vaccine had become less effective in preventing infection, and if so, in which population groups and to what degree. In order to meet this objective, we carried out three discrete analyses to answer the following questions: a) do antibody levels (IgG) of those fully vaccinated drop over time and if so, for who and how quickly, b) what is the relationship between antibody level (IgG) and subsequent PCR-confirmed infection, and c) is there a difference in PCR-confirmed infection incidence rates between those vaccinated in the initial months of the vaccination campaign to those vaccinated later?

## METHODS

We conducted a series of retrospective cohort analyses to meet the study objectives. All data were extracted from the Maccabi HealthCare Services database. Maccabi is the second largest HMO in Israel, providing healthcare coverage for over 2.5 million citizens (a quarter of Israel’s population). The database includes demographic data (date of birth, gender, socioeconomic status based on census and national survey classifications applied to home address), laboratory data (all PCR and IgG test results) and health status data (chronic illness registries, such as heart disease, HTN, CKD, diabetes and immunosuppressive disorder, based on hospital and community-based diagnoses and procedures, relevant laboratory and test results). The study was approved by the Maccabi Helsinki Committee (#0178-20-MHS), with informed consent waived, as all data extracted from the database were anomized and aggregated.

### Testing Procedures

PCR testing is carried out free of charge for any HMO member presenting with symptoms or reporting exposure to a confirmed case. Testing is carried out using real-time polymerase chain reaction tests (PCR) using Allplex™ 2019-nCoV Assay, Seegene Inc. Serology testing was carried out among specific target populations (such as employees (19%) and residents and employees of geriatric medical and retirement home facilities owned by the HMO (4%)) at discrete points in time, but the majority of testing was carried out in the general HMO population (77%) for whom testing is freely available upon request. IgG antibody testing was carried out using SARS-CoV-2 specific anti-spike antibodies with follow-up chemo-luminescence immunoassay (Quant II IgG anti-Spike CoV2-SARS by Abbott (Illinois, USA)) and reported as AU/ml (Arbitrary units). Antibody levels are reported numerically, except for outliers (below 21 and above 40,000 AU/ml) which are coded. Coded results were converted to numeric results (‘below 21’ as 21 and ‘above 40,000’ as 40,001).

### IgG antibody levels of the vaccinated population over time

All HMO members that had received both vaccine doses and had a subsequent IgG test for SARS-CoV-2 antibodies at least seven days after the second vaccination were included in this component of the study. The study period extended from 11^th^ January 2021 (when those first vaccinated reached day seven post-second dose) to 7^th^ July 2021. IgG antibody results were mapped over the 180-day period, by demographic and health characteristics (age group, gender, socio-economic status and presence of selected chronic illnesses).

### Relationship between IgG antibody levels and subsequent SARS-CoV-2 Infection

All HMO members that had a PCR test (irrespective of vaccination status) between 1^st^ June 2021 and 14^th^ July 2021 (peak of fourth wave of infection to date) with an IgG serology test between seven and 120 days prior to the PCR test were included in this component of the study. Last test result was included for those with more than one test. Last PCR test date was utilized if all results were negative and first positive PCR test date was utilized for those more than one result. The proportion of participants with subsequent positive PCR results by antibody level status was calculated.

### Comparison of infection rates by vaccination period

All HMO members who, as of the 9.6.2021, were at least seven days post-second vaccination dose with no prior positive PCR result were included in this component of the study. Members having received three doses or with an appointment to receive the third dose (N=320) in the follow-up period were excluded from analysis. (At this time, a recommendation to offer a booster vaccination for those with immune-suppressive disorder had been authorized). The study population was categorized by vaccination completion: Jan-Feb 2021 and Mar-May 2021. For both groups, the proportion that were tested PCR-positive between 9.6.2021-18.7.2021 (yes/no) was calculated.

### Statistical Analyses

Mann Whitney and Kruskal Wallis tests were carried out to compare antibody levels over time between different population groups. Linear regression was used to identify those factors associated with serology level. Ln of serology levels evidenced a normal distribution and was selected as the outcome variable. Other variables entered into a hierarchical model were a) days from vaccination, b) age, gender and socio-economic status, and c) selected chronic illnesses.

Chi square analyses were used to test the association between serology levels (categorized) with PCR outcome. Serology status was categorized into below or equal to 300 AU/ml, or above 300 AU/ml.

Kaplan-Meier survival curves were calculated to compare time from serologic test to positive PCR result for the serology categories using log-rank tests. Event was defined as positive PCR result. Time to event was the number of days from serology test to PCR test, with censoring for those that died or left the HMO, or had a follow up of less than 120 days. Logistic regression analysis was used to compare PCR-positive outcome between vaccination periods, while controlling for age group, socio-economic status and presence of chronic illness (heart disease, HTN, diabetes, CKD, and immunosuppressive disorder). Statistical analyses were carried out using SPSS, version 25 (BMI©) and R (version 3.6.2)

## RESULTS

### IgG antibody levels of the vaccinated population over time (N=8,395)

The description of the study population is provided in Table 1. Of all HMO members who received both vaccine doses, those subsequently tested for IgG antibodies were more likely to be male, younger (in the 18-44 age group), in a higher socio-economic bracket and less likely to have a chronic illness than those not IgG tested.

**Table 1:** Demographic and Health Characteristics of vaccinated population by serology test status, Jan-Jun 2021, Maccabi HealthCare Services, Israel

| Characteristic | Category | No. (%) Not Tested<br>N=1,423,257 | No. (%) Tested<br>N=8,395 |
| --- | --- | --- | --- |
| Gender | Males | 683,946 (48.1) | 2,774 (33.0) |
|  | Females | 739,311 (51.9) | 5,621 (67.0) |
| Age Group | < 18 | 39,123 (2.7) | 92 (1.1) |
|  | 18-44 | 644,038 (45.3) | 2,199 (26.2) |
|  | 45-59 | 384,100 (27.0) | 3,016 (35.9) |
|  | 60-74 | 255,377 (17.9) | 2,515 (30.0) |
|  | 75+ | 100,619 (7.1) | 573 (6.8) |
| Socioeconomic bracket | Low | 481,470 (33.8) | 3,202 (38.1) |
|  | Middle | 232,824 (16.4) | 1,281 (15.3) |
|  | High | 708,963 (49.8) | 3,912 (46.6) |
| Heart Disease | No | 1,346,990 (94.6) | 7,790 (92.8) |
|  | Yes | 76,267 (5.4) | 605 (7.2) |
| Diabetes | No | 1,297,140 (91.1) | 7,342 (87.5) |
|  | Yes | 126,117 (8.9) | 1,053 (12.5) |
| Hypertension | No | 1,145,327 (80.5) | 5,960 (71.0) |
|  | Yes | 277,930 (19.5) | 2,435 (29.0) |
| CKD | No | 1,353,406 (95.1) | 7,715 (91.9) |
|  | Yes | 69,851 (4.9) | 680 (8.1) |
| Immunosuppressive Disorder | No | 1,396,529 (98.1) | 7,411 (88.3) |
|  | Yes | 26,728 (1.9) | 984 (11.7) |

Serology levels in the study population were found to decrease over time from a mean of 14,008 for those tested within a month of being vaccinated to a mean of 1,411 for those tested in the sixth month after vaccination (Table 4). A decrease over time was observed in all sub-population groups, when results were stratified by age group, gender, socio-economic status and selected chronic illnesses (Table 2). The largest mean differences between sub-populations were observed in their initial serology levels (within the first month). Serology levels of participants aged 60 and over were almost half those of participants under the age of 60 in the first month, attenuating to a less than 10% difference six months later (Figure 1). Large differences in initial serology levels were also observed for participants with chronic illness, in particular participants with immunosuppressive disorder, CKD or heart disease. Initial (first-month) serology levels increased by socio-economic level.

**Table 2:**
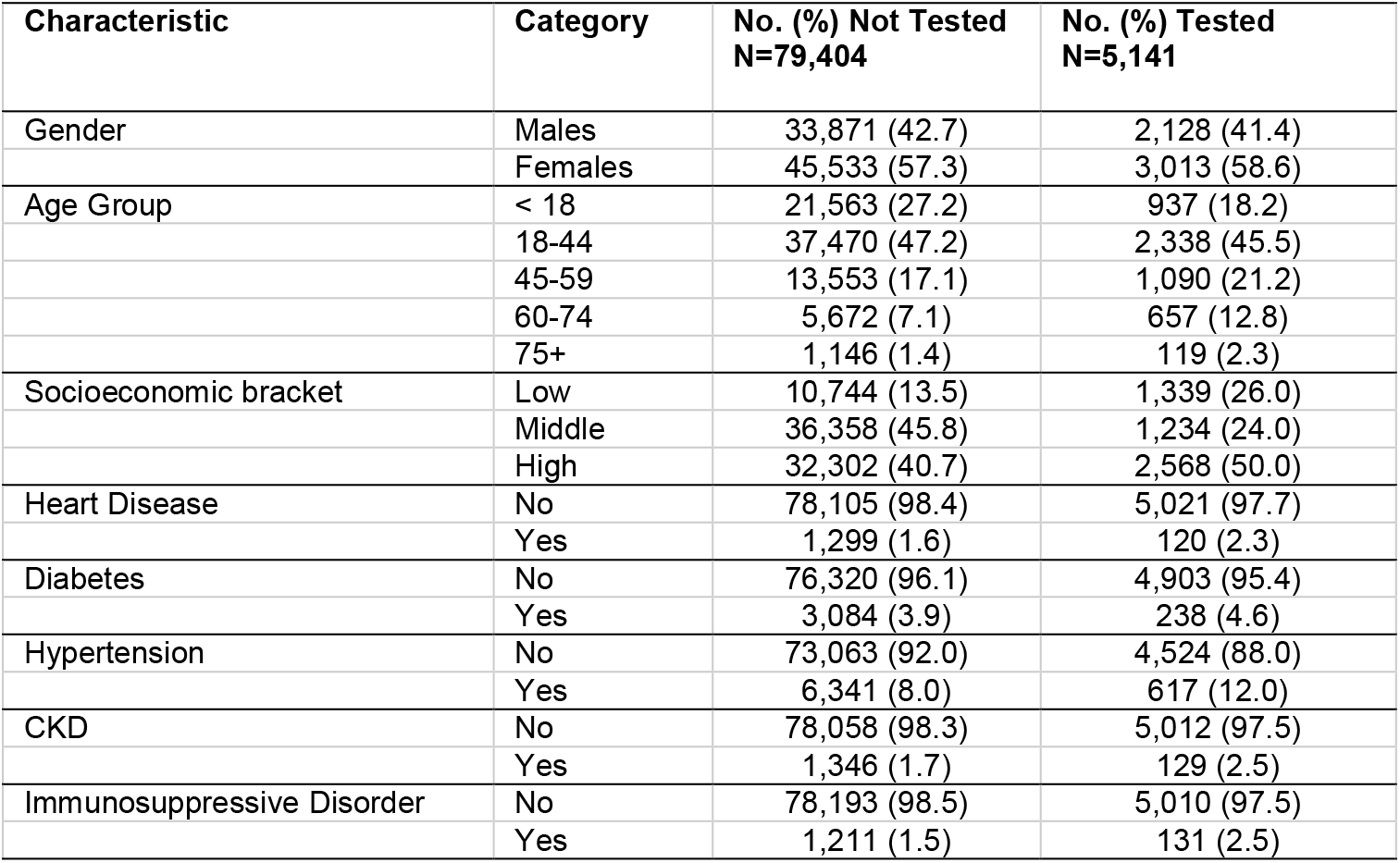
Demographic and Health Characteristics of vaccinated population by PCR test status, June-July 2021, Maccabi HealthCare Services, Israel

**Table 3:** Demographic and Health Characteristics of vaccinated population by vaccination period, Jan-May 2021, Maccabi HealthCare Services, Israel

| Characteristic | Category | Jan-Feb<br>N=821,231 | Mar-May<br>N=601,867 |
| --- | --- | --- | --- |
| Gender | Males | 394,546 (48.0) | 285,089 (47.6) |
|  | Females | 426,685 (52.0) | 313,482 (52.4) |
| Age Group | < 18 | 21,232 (2.6) | 62,793 (10.4) |
|  | 18-44 | 211,351 (25.7) | 414,514 (68.9) |
|  | 45-59 | 289,813 (35.3) | 86,013 (14.3) |
|  | 60-74 | 219,437 (26.7) | 28,620 (4.8) |
|  | 75+ | 79,398 (9.7) | 9,927 (1.6) |
| Socioeconomic bracket | Low | 102,689 (12.5) | 125,924 (20.9) |
|  | Middle | 398,474 (48.5) | 307,724 (51.1) |
|  | High | 320,068 (39.0) | 168,219 (27.9) |
| Heart Disease | No | 755,979 (92.1) | 592,608 (98.5) |
|  | Yes | 65,252 (7.9) | 9,259 (1.5) |
| Diabetes | No | 717,950 (87.4) | 581,893 (96.7) |
|  | Yes | 103,281 (12.6) | 19,974 (3.3) |
| Hypertension | No | 593,368 (72.3) | 556,545 (92.5) |
|  | Yes | 227,863 (27.7) | 45,322 (7.5) |
| CKD | No | 762,717 (92.9) | 592,447 (98.4) |
|  | Yes | 58,514 (7.1) | 9,420 (1.6) |
| Immunosuppressive Disorder | No | 808,061 (98.4) | 598,499 (99.4) |
|  | Yes | 13,170 (1.6) | 3,368 (0.6) |

**Table 4:**
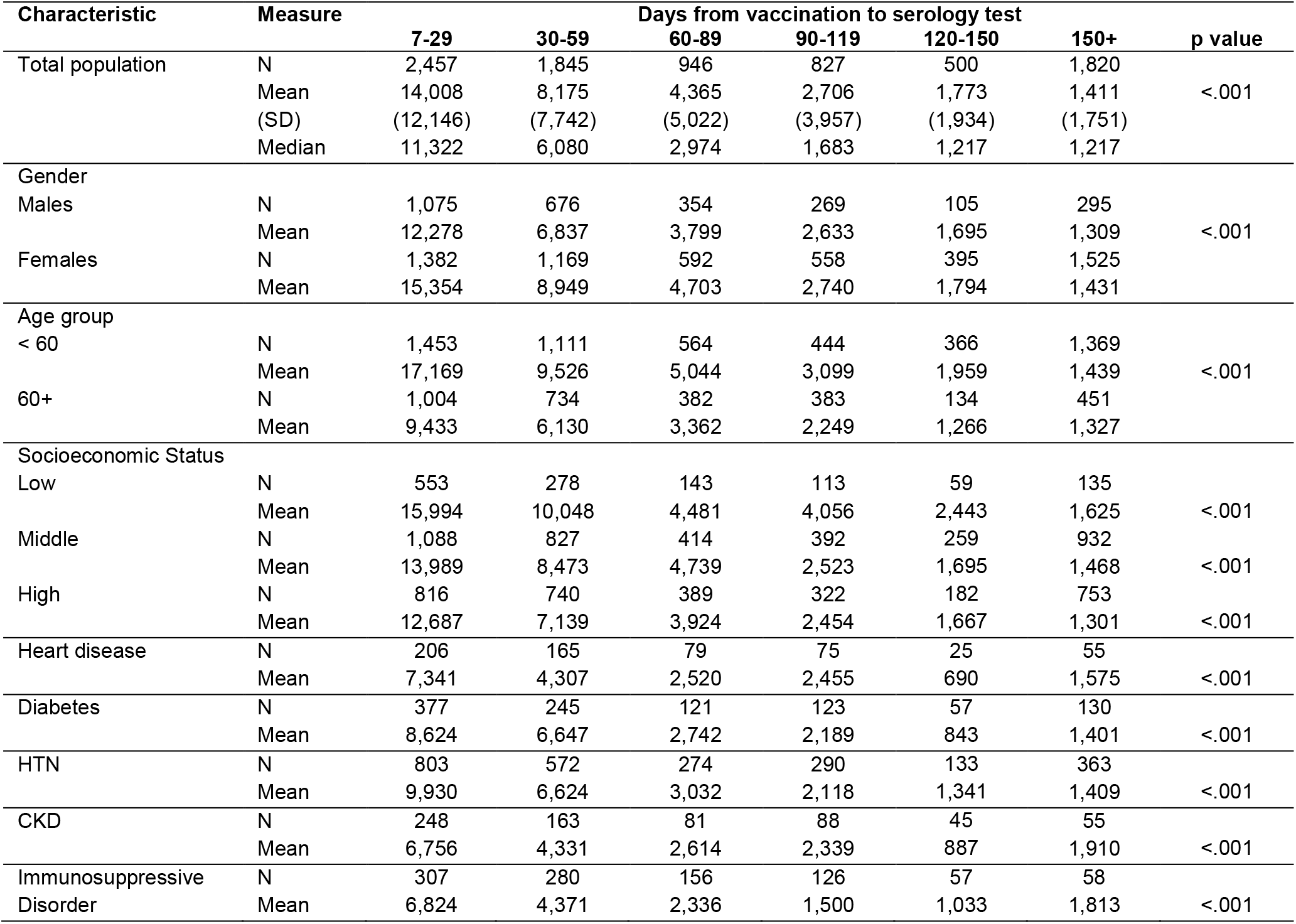
Mean antibody level by demographic and health variables and time from vaccination (days), Jan-June 2021, Maccabi HealthCare Services, Israel

**Figure 1:**
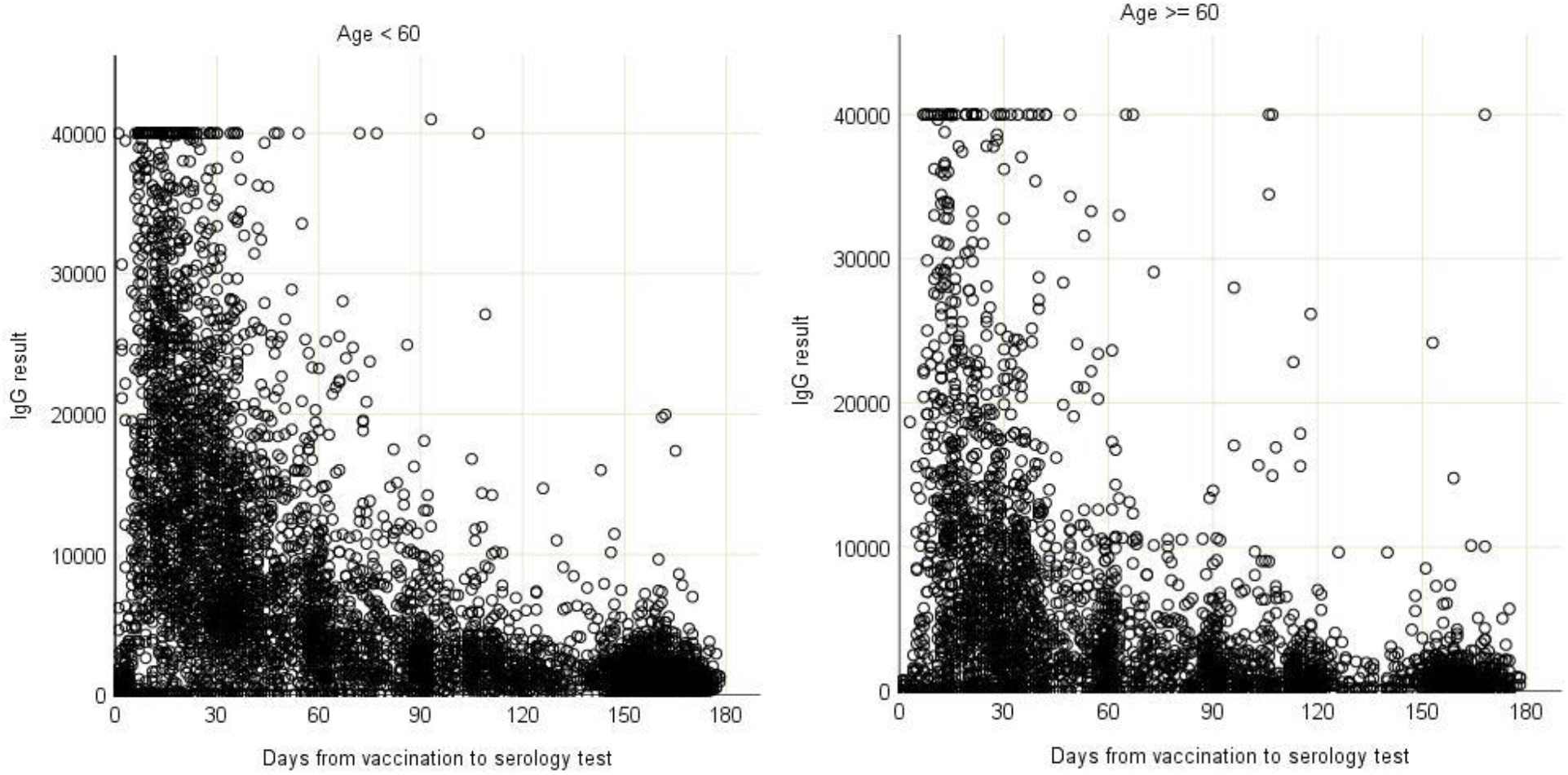
Serology levels of vaccinated population over time by age group, Jan-June 2021, Maccabi HealthCare Services, Israel

Of all those that were vaccinated with both doses, 2.8% also had a positive PCR result. Comparable declines in serology means by month were observed in this group, as in others. However, the mean serology level for those tested in the first 7-30 days was much higher than for the total study population (22,630 AU/ml, p<.001).

When demographic and health variables were entered into a linear regression model (Appendix, Table A1), all factors remained independently associated with serology levels, with the highest co-efficient observed for participants with immunosuppressive disorder. No multi-collinearity was observed between the factors in the regression model.

### Relationship between IgG antibody levels and subsequent SARS-CoV-2 Infection (N=5,141)

Demographic and health characteristics of HMO members with a serology result who had subsequently tested for PCR were similar to those that had no PCR test result (Table 2) with the exception of socio-economic status and diabetes status. PCR testers were more likely to be in the lower socio-economic bracket and have diabetes than non-PCR testers.

Of all those who had both tests (N=5,141), 57% had a serology test result of 150 AU/ml and lower, 6% had a result between 150-299 AU/ml, 10% had a test result between 300-799 AU/ml and 27% had a result greater than 800 AU/ml. The proportion of participants with a positive PCR result were 1.2% for those with serology levels lower 150 AU/ml and lower, 1.3% for those with serology levels between 150-299, 0.2% of those with a result of 300-799 AU/ml and for those with a result of 800+ AU/ml (p=.004). Mean serology levels for the 42 study participants with a positive PCR result was 175 (SD: 490) compared to a mean serology level of 2057 (SD: 6030) for those with a negative result (p<.001).

Of all the study participants for this component of the study, 365 (7%) had a prior infection (37% of whom received one vaccine dose). Those with a prior infection were less likely to have a serology level of 300 AU/ml and below than those without a prior infection (40.3% vs 65.2% respectively, p<.001.

However, irrespective of prior infection (yes/no), the proportion of those with a new PCR positive result was 0.8%. Kaplan-Meier survival curves (over time by serology status (+/- 300 AU/ml) are presented in Figure 2. The curves indicate that participants with lower serology level (</=300 AU/ml) had lower survival rates than participants with higher serology status ((+/- 300 AU/ml) (log-rank test p=.003).

**Figure 2:**
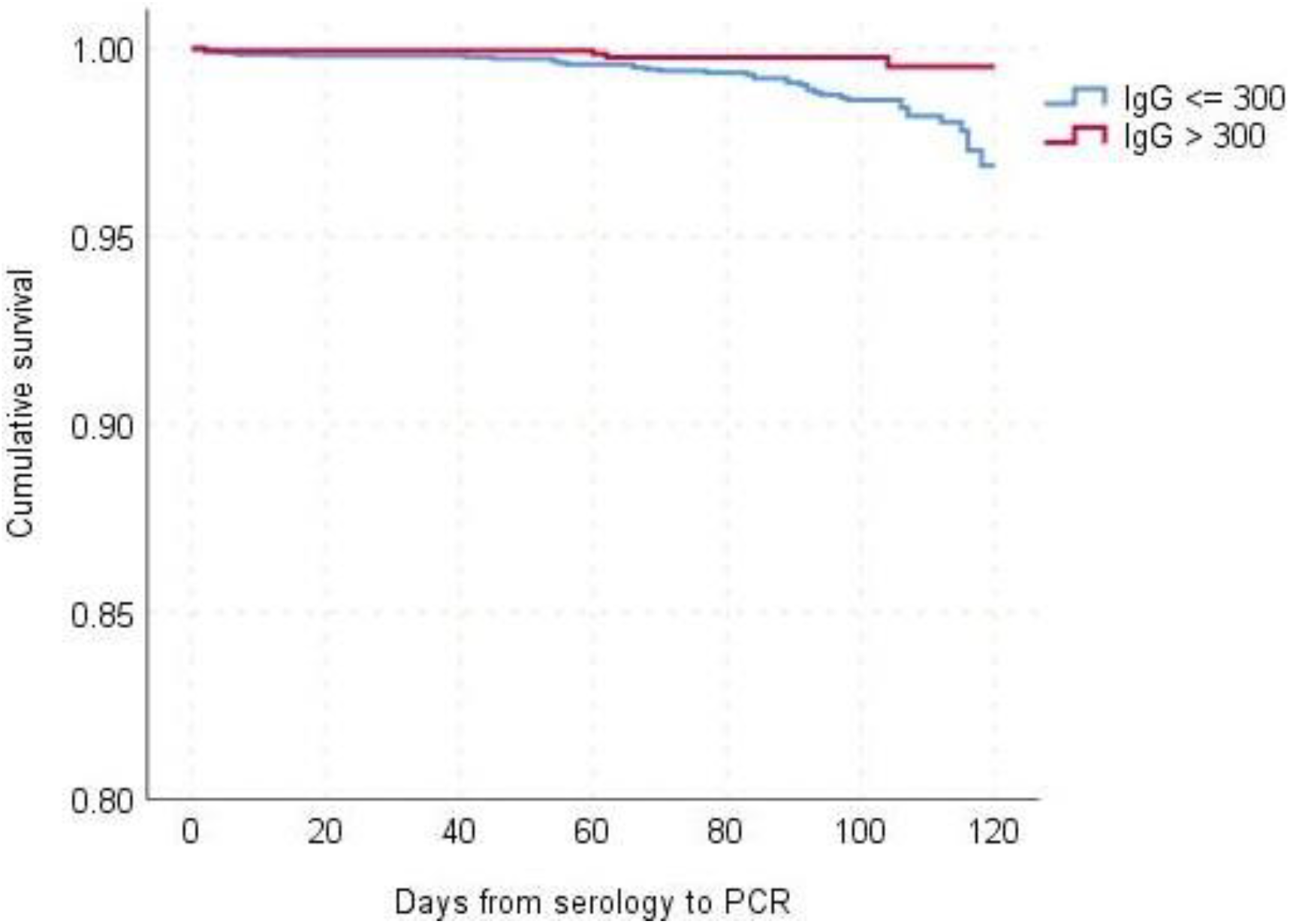
Kaplan-Meier cumulative survival for PCR positive outcome, June-July 2021, Maccabi HealthCare Services, Israel

### *Comparison of infection rates by vaccination period* (N=1,423,098)

At the time of the study, 86% of those eligible for vaccination (aged 16+) had received both doses in the HMO (90% of those aged 60+). Comparison of demographic and health variables between those vaccinated in the first two months, compared to those vaccinated later are presented in Table 3. Those vaccinating in the first two months were more likely to be older, in a higher socio-economic bracket and with higher rates of chronic illness. 1,518 (0.19%) of those vaccinated in Jan-Feb 2021 were subsequently found to be PCR positive, compared to 644 (0.11%) of those vaccinated in Mar-May 2021 (p<.001). Univariate analyses (Appendix, Table A2) also revealed that age, gender, socio-economic status and presence of chronic illnesses (health disease, diabetes, HTN and CKD) were all associated with having a positive PCR result.

Factors associated with subsequent infection (positive PCR result) in a logistic regression model (Table 5) were socio-economic status, age group, vaccination period, gender and heart disease. When controlling for all other factors, members vaccinated first were 1.6 times more likely to get infected with CoVid-19 than those vaccinated later.

**Table 5:** Factors associated with Covid-19 infection (logistic regression model), Jan-May 2021, Maccabi HealthCare Services, Israel

| Variable | Category | N | OR | 95% CI |
| --- | --- | --- | --- | --- |
| Vaccination period | Jan-Feb | 821,231 | 1.61 | 1.45-1.79 |
|  | Mar-May | 601,867 | - |  |
| Gender | Males | 681,382 | 0.11 | 1.01-1.20 |
|  | Females | 741,716 | - |  |
| Age Group | < 18 | 84,025 | - |  |
|  | 18-44 | 625,865 | 1.92 | 1.51-2.49 |
|  | 45-59 | 375,826 | 1.88 | 1.46-2.45 |
|  | 60-74 | 248,057 | 1.54 | 1.17-2.04 |
|  | 75+ | 89,325 | 1.06 | 0.75-1.50 |
| Socioeconomic | Low | 228,613 | - |  |
|  | Moderate | 706,198 | 2.85 | 2.34-3.50 |
|  | High | 488,287 | 4.40 | 3.61-5.41 |
| Heart Disease | No | 1,348,587 | - |  |
|  | Yes | 74,511 | 1.35 | 1.11-1.79 |
| Diabetes | No | 1,299,843 | - |  |
|  | Yes | 123,255 | 1.03 | 0.87-1.12 |
| Hypertension | No | 1,149,913 | - |  |
|  | Yes | 273,185 | 0.98 | 0.86-1.22 |
| CKD | No | 1,355,164 | - |  |
|  | Yes | 67,934 | 0.82 | 0.63-1.05 |
| Immunosuppressive Disorder | No | 1,406,596 | - |  |
|  | Yes | 16,502 | 0.90 | 0.58-1.63 |

## DISCUSSION

In this study, we established that IgG serology levels for SARS-CoV-2 virus decreased progressively over time for both the total vaccinated population and in each sub-population when stratified by demographic and health variables. We also established a clear association between serology levels and subsequent risk of infection, wherein participants with a serology level of 300 AU/ml or lower were more likely to get infected with CoVid-19 than those with a serology level above 300 AU/ml. Finally, we established that those vaccinated at the beginning of the national vaccination campaign were more likely to get infected (during the current wave of infection) than those vaccinated later. These findings suggest that effectiveness of the vaccine does drop over time and that the current wave of infection can be attributed, at least in part, to the vaccine’s reduced effectiveness over time.

### IgG antibody levels of the vaccinated population over time

Initial serological studies focused on patients found PCR positive for CoVid-19, and reported a decrease over time of antibody presence from time of infection.*(9-11)* Fewer studies have looked specifically at serological response of the vaccinated population. Most of the studies based on vaccinated populations found near 100% seroconversion rates, but had short follow-up periods.*(12,13)* Serological levels were much higher amongst the vaccinated population than those convalescing after infection*(12)* and those under the age of 50.*(13)* In a case-control study of PCR positive cases divided by prior vaccination status (yes/no), Lopez-Bernal et al. (2021)*(8)* found that those vaccinated (two dose) with the BNT162b2 vaccine with the Alpha variant achieved 93.7% vaccine effectiveness, compared to an 88% vaccine effectiveness rate for those infected with the Delta variant.

We did not find studies published describing serological status over longer follow-up periods for a vaccinated population. Mean levels of IgG antibody decreased progressively over time for all sub-populations in this study. The difference between the groups was mostly evident in initial (first month) starting means, with the elderly and those suffering from chronic illness having lower levels, but attenuating to more comparable levels between groups six months after vaccination. In a large British household study, IgG response was measured over the first three months*(14)* found higher sero-conversion rates for younger age groups (20-40), females, those receiving both doses, vaccination with BNT162b2 vaccine (compared to Astra-Zeneca vaccine) and those with evidence of a prior infection. Low responders were older and had higher prevalence of chronic illness/disease, such as patients on immune-suppressants or suffering from diabetes. These same population groups, were found in this study to start with lower serology levels and have lower mean serology levels six-months post-vaccination.

### Relationship between IgG antibody levels and subsequent SARS-CoV-2 Infection

One of the many unknowns regarding CoVid-19 is to what extent IgG antibody presence is indicative of protection against the virus. The manufacturer’s recommended cut-off indicating a positive serological response (<50 AU/ml) is much lower than the mean serology levels found in the sixth month post-vaccination in the present study. Are higher levels indicative of higher protection? Other mechanisms of protection, such as anti-viral T and B cell memory have been suggested as offering protection, even in the absence of seroconversion.*(15)* Khoury et al (2021)*(16)*, in a meta-analysis, found a strong relationship between mean neutralization levels and reported protection. They further estimated that protection was likely to fall over 250 days, although with still largely preserved protection from severe infection. In the second component of our study, we found a strong association between serological level and PCR outcome where increasing serology level was associated with decreasing infection rates. Using a cut-off of 300 AU/ml, we found higher rates of infection for those with low serology levels.

### Comparison of infection rates by vaccination period

Little data is available to compare vaccine effectiveness over time, with observational follow-up studies becoming less appropriate, given the potential bias between those electing to vaccinate and those that do not. Pfizer-BionTech published its most recent efficacy study, wherein symptomatic infection rates were compared between vaccinated and unvaccinated groups over a period of six months.*(7)* Vaccine efficacy for infection dropped from 96% within the first two months post-second dose to 84% vaccine efficacy four to six months post-second dose. Consistent with Pfizer’s findings, we found higher rates of CoVid-19 infection among those vaccinated in the initial months of the vaccine campaign compared to those vaccinated later. Even after controlling for age (those vaccinated first were more likely to be older), incidence rates were higher in the first-vaccinated group. Were the majority of the current wave of infection be attributable to the Delta virus, we would expect consistent incidence rates, irrespective of when the individual was vaccinated. We suggest that the difference found here between time periods indicates a reduction in vaccine effectiveness over time. We cannot rule out, however, some contribution of the Delta variant to reduced effectiveness.

The study findings here are subject to a number of limitations. Firstly, serology test findings were not based on repeated tests in the same population, but a description of the results over time of a non-randomly selected population. Those presenting for serology and PCR testing were not a randomly selected group. Mean serology levels were calculated for each sub-population, despite the potential for outlier measures to skew results, to allow statistical comparison between sub-population groups. Numbers were small for some stratified data, particularly for the 120-149 period, and should be interpreted with caution. Study findings were not adjusted for serology test accuracy. Conclusions are made on the assumption that the majority of those infected in the third component of the study (by time of vaccination) were infected with the Delta variant, given its’ prevalence in Israel.

Given these limitations, the different elements of the study were based on large numbers of a vaccinated population with six months of follow-up time to measure CoVid-19 infection. We established that serology levels for all groups have decreased over time, that there is a strong relationship between serology levels and subsequent infection, and that those vaccinated early in the vaccination campaign had higher infection rates. These factors taken together suggest that the BNT162b2 vaccine, as indicated by the manufacturer, offers lower protection against infection over time, independent of variant type. These results, combined with other studies carried out in Israel and elsewhere, contributed to the decision to offer a third dose of the BNT162b2 vaccine to the population aged 60 and over. Follow-up of infection and morbidity rates in this group will allow us to confirm the wisdom of providing a booster dose.

## Data Availability

Available from corresponding author upon reasonable request

## Acknowledgments

none

## Disclaimers

The opinions expressed by authors contributing to this journal do not necessarily reflect the opinions of the Centers for Disease Control and Prevention or the institutions with which the authors are affiliated.” Additional disclaimers are discouraged.

## Author Bio

Jennifer Kertes MPH, is a senior researcher in the Division of data & digital health, Maccabi Healthcare Services and has been involved in the design and implementation (data extraction, analysis and interpretation and summary) of innumerable epidemiological and public health studies for the HMO for over twenty years. Her primary research interests include public health and health promotion studies (especially regarding smoking) and pharma-therapeutic studies.

## Appendix

**Table A1:** Factors associated with ln of serology titers (linear regression model)

| Factor | $\beta$ | p value |
| --- | --- | --- |
| Days from vaccination | -.016 | <.0001 |
| Gender (males) | -.347 | <.0001 |
| Age (years) | -.022 | <.0001 |
| Socio-economic status (middle) | -.078 | .033 |
| Socio-economic status (high) | -.184 | <.0001 |
| Heart Disease | -.242 | <.0001 |
| Diabetes | -.108 | .007 |
| HTN | -.105 | .001 |
| CKD | -.318 | <.0001 |
| Immunosuppressive disorder | -.888 | <.0001 |

$R^2 = .498$ . Dependent variable: ln of serology titer. Independent variables entered in hierarchical model: days from vaccination; gender, age and socio-economic status; chronic illnesses.

**Table A2:**
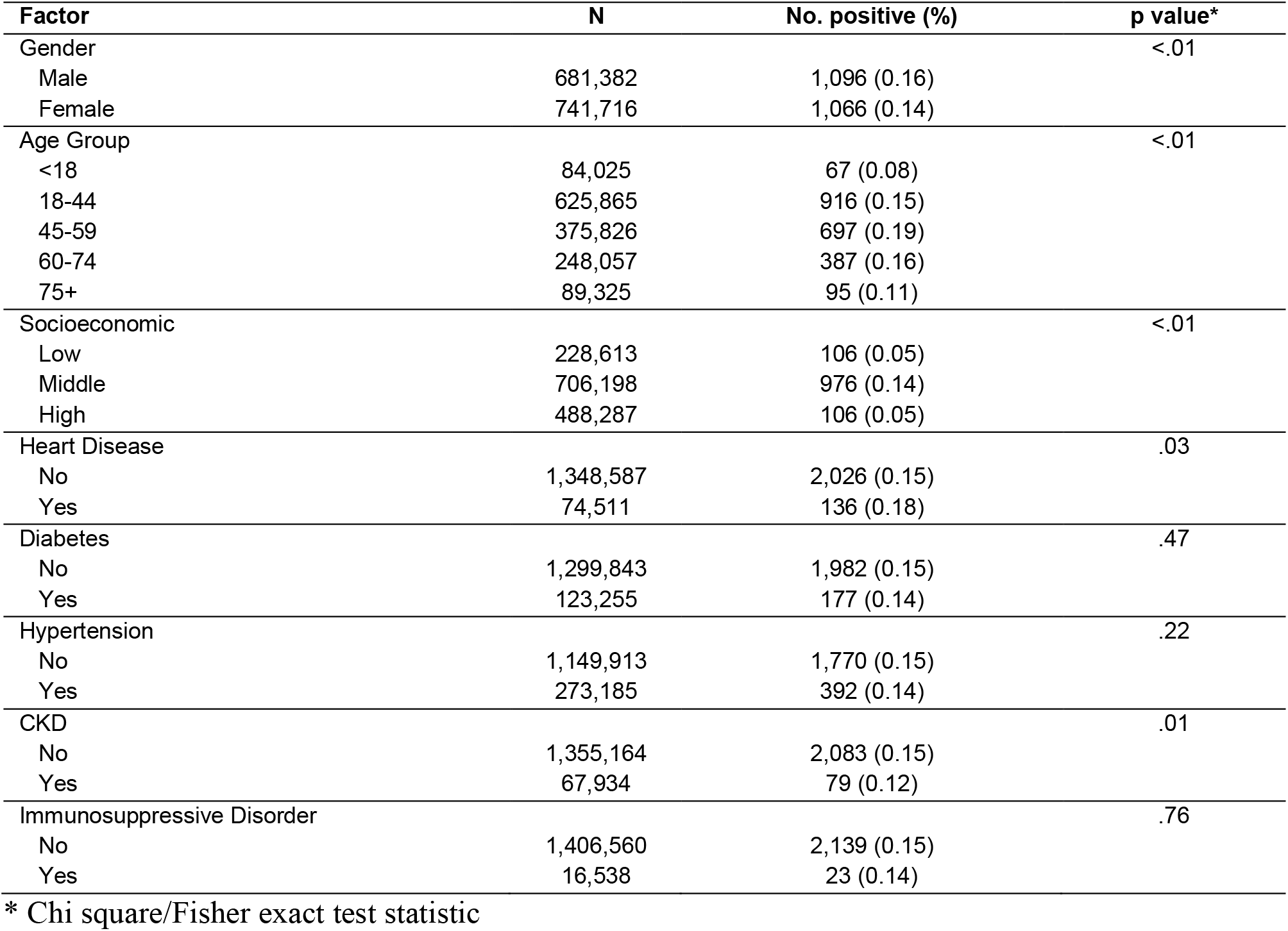
Factors associated with a positive PCR result (univariate analysis)

## References

1. Rosen B, Waitzberg R, Israeli A. Israel’s rapid rollout of vaccinations for COVID-19. Isr J Health Policy Res. 2021;10(1):1–14.

2. Leshem E, Wilder-Smith A. COVID-19 vaccine impact in Israel and a way out of the pandemic. Lancet. 2021;397(10287):1783–1785.

3. Vaccines and Related Biological Products Advisory Committee Meeting, Dec 10<sup>th</sup> 2020. FDA Briefing Document - Pfizer BioNTech COVID-19 vaccine. 2021. https://www.fda.gov/media/144245/download

4. Polack FP, Thomas SJ, Kitchin N, Absalon J, Gurtman A, Lockhart S, et al. Safety and efficacy of the BNT162b2 mRNA Covid-19 vaccine. N Engl J Med. 2020;382(27):2603–2615.

5. Haas EJ, Angulo FJ, McLaughlin JM, Anis E, Singer SR, Khan F, et al. Impact and effectiveness of mRNA BNT162b2 vaccine against SARS-CoV-2 infections and COVID-19 cases, hospitalisations, and dealth following a nationwide vaccination campaign in Israel: an observational study using national surveillance data. Lancet. 2021;397(10287):1819–1829.

6. Dagan N, Barda N, Kepten E, Miron E, Perchik S, Katz MA, et al. BNT162b2 mRNA COVID-19 vaccine in a nationwide mass vaccination setting. N Engl J Med. 2020;384(15):1412–1423.

7. Thomas SJ, Moreira ED, Kitchin N, Absalon J, Gurtman A, Lockhart S, et al. Six month safety and efficacy of the BNT162b2 mRNA COVID-19 vaccine. medRxiv [Preprint] [Cited July 28, 2021]. Available from https://doi.org/10.1101/2021.07.28.21261159

8. Lopez Bernal J, Andrews N, Gower C, Gallagher E, Simmons R, Thelwall S, et al. Effectiveness of Covid-19 vaccines against the B.1.617.2 (Delta) Variant. N Engl J Med. Aug 12;385(7):585–594. Epub 2021 Jul 21.

9. Zhao J, Yuan Q, Wang H, Liu W, Liao X, Su Y, et al. Antibody responses to SARS-CoV-2 in patients of novel coronavirus disease 2019. Clin Infect Dis. 2020;71(16):2027–2034.

10. Zhou W, Xu X, Chang Z, Wang H, Zhong X, Tong X, et al. The dynamic changes of serum IgM and IgG against SARS-CoV-2 in patients with COVID-19. J Med Virol. 2021;93(2):924–933.

11. Harvey RA, Rassen JA, Kabelac CA, Turenne W, Leonard S, Klesh R, et al. Association of SARS-CoV-2 seropositive antibody test with risk of future infection. JAMA Intern Med. 2021;181(5):672–679.

12. Jalkanen P,, Kolehmainen P, Häkkinen HK, Huttunen M, Tähtinen PA, Lundberg R, et al. COVID-19 mRNA vaccine induced antibody responses against three SARS-CoV-2 variants. Nat Commun. 2021;12(1):3991.

13. Grupel D, Gazit S, Schreiber L, Nadler V, Wolf T, Lazar R, et al. Kinetics of SARS-CoV-2 anti-S IgG after BNT162b2 vaccination. Vaccine. Aug 11;S0264-410X(21)01045-8. Epub PMID: 34393018

14. Wei J, Stoesser N, Matthews PC, Ayoubkhani D, Studley R, Bell I, et al. Antibody responses to SARS-CoV-2 vaccines in 45,965 adults from the general population of the United Kingdom. Nat Microbiol. July 2021:1–10. Epub. PMID: 34290390

15. Cox RJ, Brokstad KA. Not just antibodies: B cells and Tcells mediate immunity to COVID-19. Nat Rev Immunol. 2020;20(10):581–582.

16. Khoury DS, Cromer D, Reynaldi A, Schlub TE, Wheatley AK, Juno JA et al. Neutralizing antibody levels are highly predictive of immune protection from symptomatic SARS-CoV-2 infection. Nat Med. 2021;27(7):1205–1211.

